# Comparative Analysis of Early Dynamic Trends in Novel Coronavirus Outbreak: A Modeling Framework

**DOI:** 10.1101/2020.02.21.20026468

**Authors:** Huazhen Lin, Wei Liu, Hong Gao, Jinyu Nie, Qiao Fan

**Affiliations:** Center of Statistical Research and School of Statistics, Southwestern University of Finance and Economics, Chengdu, China; Centre for Quantitative Medicine, Program in Health Services & Systems Research, Duke-NUS Medical School, Singapore

**Author notes:** Email addresses:* (Huazhen Lin), (Qiao Fan).

**Keywords:** Coronavirus, COVID-19, SARS-CoV-2, transmissibility, outbreaks, reproduction number *R*, generalized growth modeling

## Abstract

**Background:** The 2019 coronavirus disease (COVID-19) represents a significant public health threat globally. Here we describe efforts to compare epidemic growth, size and peaking time for countries in Asia, Europe, North America, South America and Australia in the early epidemic phase.

**Methods:** Using the time series of cases reported from January 20, 2020 to February 13, 2020 and transportation data from December 1, 2019 to January 23, 2020 we have built a novel time-varying growth model to predict the epidemic trend in China. We extended our method, using cases reported from January 26, 2020 - or the date of the earliest case reported, to April 9, 2020 to predict future epidemic trend and size in 41 countries. We estimated the impact of control measures on the epidemic trend.

**Results:** Our time-varying growth model yielded high concordance in the predicted epidemic size and trend with the observed figures in C hina. Among the other 41 countries, the peak time has been observed in 28 countries before or around April 9, 2020; the peak date and epidemic size were highly consistent with our estimates. We predicted the remaining countries would peak in April or May 2020, except India in July and Pakistan in August. The epidemic trajectory would reach the plateau in May or June for the majority of countries in the current wave. Countries that could emerge to be new epidemic centers are India, Pakistan, Brazil, Mexico, and Russia with a prediction of 10^5^ cases for these countries. The effective reproduction number *R*_*t*_ displayed a downward trend with time across countries, revealing the impact of the intervention remeasures i.e. social distancing. *R*_*t*_ remained the highest in the UK (median 2.62) and the US (median 2.19) in the fourth week after the epidemic onset.

**Conclusions:** New epidemic centers are expected to continue to emerge across the whole world. Greater challenges such as those in the healthcare system would be faced by developing countries in hotspots. A domestic approach to curb the pandemic must align with joint international efforts to effectively control the spread of COVID-19. Our model promotes a reliable transmissibility characterization and epidemic forecasting using the incidence of cases in the early epidemic phase.

## 1. Introduction

In early December 2019, a novel severe acute respiratory syndrome coronavirus, SARS-COV-2, emerged into the human population in Wuhan, China^1–7^. The first coronavirus disease 2019 (COVID-19) case outside China was reported on January 13, 2020 in Thailand. In just several weeks, the local transmission started rapidly in a broad array of countries in Asia, Europe, North America, South America and Australia, with the emergence of new epicenters of spread, such as the US, Italy, Spain, and France (https://www.who.int). The rapid spreading of SARS-COV-2 has led to a major global public health threat. The World Health Organization (WHO) declared the spread of COVID-19 a pandemic on March 11, 2020.

Early epidemic forecasts consisting of the likely trajectory of an unfolding outbreak can help guide the type and intensity of interventions^8–10^. The vast majority of these approaches considered early exponential growth dynamics, an assumption that could lead to substantial overestimation of epidemic size and peak timing. To enhance the ability to forecast epidemics, it is crucial to characterize the shape of epidemic growth and accurately assess early trends of sub-exponential growth phenomenon^11–13^. Currently, the global case count continues to rise, but there is a limited understanding of the extent of the outbreak and epidemic growth profile, particularly for the new emerging epicenters.

In this study, we attempt to assess and compare the extent of the outbreak across countries, draw preliminary conclusions about the impact of control measures, and characterize real-time effective reproduction number *R*_*t*_. Our model, without making explicit assumptions about the epidemic growth profile, is a generalizable framework to estimate the early dynamic trends of COVID-19 from the incidences of cases. We estimated the epidemic trend and size in China using the cases reported from January 20, 2020 to February 13, 2020, and in other 41 countries using data from January 26, 2020, - or the date of the earliest case reported, to April 9, 2020.

## 2. Methods

### 2.1. Sources of Data

We obtained the number of COVID-19 confirmed cases of time series data between January 20, 2020 to February 13, 2020 in China from the official websites of the National Health Commission of China and Provincial Health Committees (http://www.nhc.gov.cn). The data of cases for each of the 29 provinces (25 provinces plus 4 municipalities including Beijing, Shanghai, Chongqing and Tianjin) at 23 time points were included, as well as for Wuhan and major cities in Hubei. The start date of January 20 was chosen because the official diagnostic protocol released by WHO on January 17 allows the new COVID-19 cases to be diagnosed accurately and rapidly. All cases were laboratory-confirmed with the detection of viral nucleic acid following the case definition by the National Health Commission of China.

Wuhan is connected to other cities in China via high-speed railway, highway, and airplane flights. Population mobility statistics to estimate the exposed sizes in cities outside Wuhan were based on transport-related databases below: 1) Railway and airline travel data: the daily numbers of outbound high-speed trains from Wuhan with corresponding travelling hours were obtained from the high-speed rail network (http://shike.gaotie.cn) from December 1, 2019 to January 23, 2020, and similarly daily numbers of outbound flight and hours for air transport were obtained from the Citytrip network (https://www.ctrip.com) from December 1, 2019 to January 23, 2020. We calculated daily travelling hours which equal to the product of the outbound trip counts and the travelling hours for rail and air transport respectively from Wuhan to each major city. For a given province, we summarized the total number of travelling hours across all cities in that province. 2) Highway mileage data: we collected highway mileage data from bus station networks at https://www.qichezhan.cn. It contains the highway mileage from Wuhan to 16 cities in the Hubei Province. 3) Migration data: we obtained population migration data from the Baidu Migration Map (http://qianxi.baidu.com) which includes both the percentages and volumes of migration from Wuhan to other cities and provinces from January 1 to 28, 2020. Total travelling hours for rail and air flight, and migration scales are plotted by the province in Supplementary Figure 1. Accumulated time on trains, on airplanes, highway mileage and population migration scales were used to model the underlying epidemic sizes in the provinces or cities outside Wuhan at the time 0 of this study which is on January 20, 2020.

From Supplementary Figure 1, we observed that Guangdong has the largest traveling hours through railway and airplane outbound from Wuhan among the provinces. Also, the largest population has immigrated from Wuhan to Henan. In Hubei province, Cities of Huanggang and Xiaogan are the closest to Wuhan in terms of mileage and the scale of migration. These simple observations are consistent with our result that Guangdong, Henan, Huanggang and Xiaogan have the largest number of estimated primary infected cases imported from Wuhan on January 20, 2020.

We obtained the number of COVID-19 confirmed cases in 41 countries using data from January 23, 2020, or the date of the earliest case reported, to April 9, 2020 from the offical websites of WHO at https://www.who.int. We included 41 countries with at least confirmed cases on April 9 2020 in five continents: Asia, Europe, North America, South America, and Australia.

### 2.2. Modelling the transmissibility of COVID-19

We introduce the main notation here. All times are calendar times, measured in days since the start of the epidemic.

*Y*_*kt*_ number of accumulated diagnosed case till day *t*,

*TR*_*k*_ daily traveling hours on trains from Wuhan,

*FL*_*k*_ daily traveling hours on airplane from Wuhan,

*RM*_*k*_ highway mileage from Wuhan,

*MI*_*k*_ volumes of migration from Wuhan from January 1 to 28, 2020,

*α*_*k*_ number of underlying primary infected cases,

*W*_*kt*_ underline number of infected individuals who are infectious,

*m* duration of infectious period (day),

where the subscript *k* represents country, province or city *k*, the subscript *t* represents day *t. TR*_*k*_ and *FL*_*k*_ are constructed based on the two reasons. One is that the longer people stay on the train or plane, the more likely he(she) is to get infected. Another is that the infection happens in local area, hence the number of trains or planes has more information than the population taking trains or planes. In addition, in Hubei province, most people left Wuhan by cars, we use *RM*_*k*_ as one of measurement for the spatial distance between city *k* and Wuhan.

#### 2.2.1. Modeling for 28 provinces

First, we build an index *α*_*k*_ to represent the baseline infected cases in province *k* on 20 January, 2020. Particularly, we will use *TR*_*k*_, *FL*_*k*_ and *MI*_*k*_ to measure the relationship between provice *k* and Wuhan. We suppose

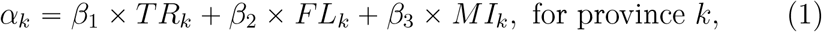

where *β* = (*β*_1_, *β*_2_, *β*_3_)′ are estimated by the observed *Y*_*kt*_ in provinces *k* = 1, …, *K* and *t* = 1, …, *T*.

So far, we are not sure the key epidemiological parameters that affected spread and persistence. We hence make assumptions as least as possible. It is obvious that the average cases in province *k* diagnosed at day *t* is proportional to the scale of infectious cases on day *t* − 1, *W*_*k,t* − 1_. We then assume a Poisson distribution for the new cases diagnosed in province *k* at day *t* with mean *γ*_*kt*_*W*_*k,t* − 1_, that is

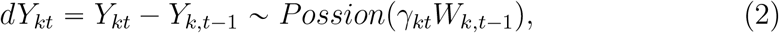

where ‘∼’ means ‘distributed as’.

Under the unified leadership of the central goverment, we suppose the trend of *γ*_*kt*_ over day *t* is the same for 28 provinces, that is, *η*_*kt*_ = *η*_*t*_ is independent of *k* so that

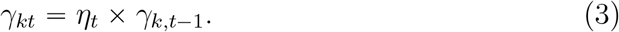

To avoid strong assumptions about the evolution of the epidemic, we allow *η*_*t*_ to be arbitray function of *t*. We determine the functional form of *η*_*t*_ by pointwise estimating *η*_*t*_ and checking the resulting pattern over *t*. Denote the resulting functional form for *η*_*t*_ by *η*_*t*_ = *η*_*t*_(*a*).

The new cases diagnosed at day *t* may be not reported fully. That is, *E*(*dY*_*kt*_) = *pdW*_*kt*_ and *p* < 1. The estimation for *p* need more information except that we have. Fortunately, simple mathematical derivation shows that the value of *p* may influence the prediction of the absolute epidemic size but will not affect the trend of the epdimic, for example, the reproduction number, the duration and the peak time of the epidemic and relative epidemic size, in which we are interested. Hence, we suppose *p* = 1 in the paper. Since

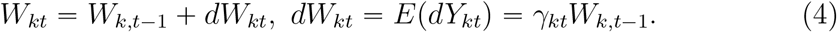

With the chain calculation, we have 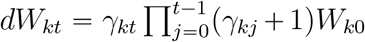.and 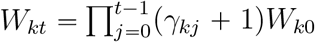,where *W*_*k0*_ = *α*_*k*_ and *γ*_*k*0_ = 0. In practice, the infected patients will be isolated and removed from the infectious source. With the notation *m* of duration of infectious period, we hence have

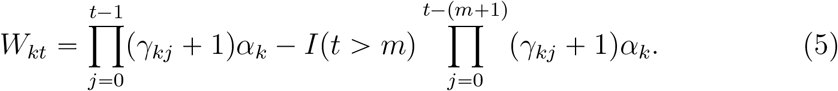

Denote *γ*_1_ = (*γ*_11_, …, *γ*_*K*1_)′ and all of the parameters by 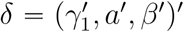. Taken (1), (2), (3) and (5) together, the loglikelihood function was

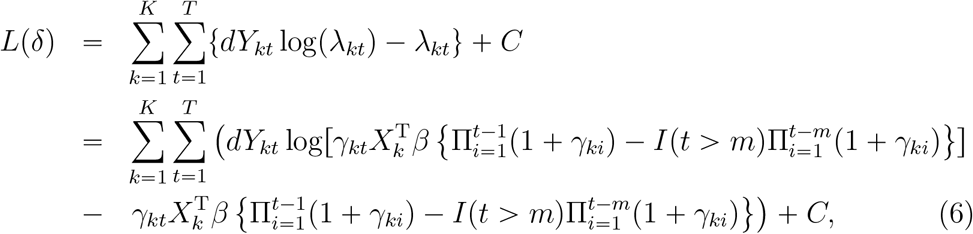

where *C* is a constant independent of *δ, m* is determined by minimizing the prediction error. The confidence intervals were obtained based on 200 bootstrap resampling^14, 15^.

#### 2.2.2. Modeling for Hubei, Wuhan and the other countries

The modeling and the loglikelihood function for Hubei are similar with those for 28 provinces except that *FL*_*k*_ is replaced by *RM*_*k*_ and provinces are replaced by cities, because there are not flights between the cities in Hubei and Wuhan, and the most people leave Wuhan by cars or buses. Specifically,

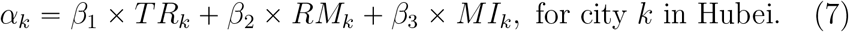

The modeling and the likelihood function for Wuhan and countries, including Singapore, South Korea, Japan, Italy, German, Span, France and Iran, are similar with those for 28 provinces except that *α*_*k*_ is directly estimated by the diagnosed cases in Wuhan and other 41 countries respectively.

#### 2.2.3. The calculation of the time-dependent reproduction number R_t_

The equation *dW*_*kt*_ = *γ*_*kt*_*W*_*k,t−*1_ implies that, when *W*_*k,t−*1_ = *1* we have *γ*_*kt*_ = *dW*_*kt*_. Hence *γ*_*kt*_ is the average number of new infections created by an infectious individual in one day, then 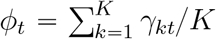 is the corresponding average number across provinces or cities. Since one infectious individual can make infection for *m* days, an infectious individual then can lead to *R*_*t*_ *mϕ*_*t*_ new infections, which indeed is the time-dependent reproduction number^16, 17^.

#### 2.2.4. Predication of potential peak time and turning point in COVID-19 outbreak

With the estimated parameters by maximizing the loglikehood (6), we can estimate and predict the average new cases 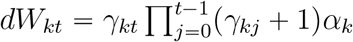, then the cumulative cases 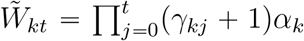. Based on the new cases and cumulative cases, we can predict the peak time and the turning point in the COVID-19 outbreak. In the paper, we defined the peak time to be the day at which the incidence cases began to decline, and the turning point to be the day when the number of the cumulative cases reached a plateau, which satisfying ∣*∂f*(*v*)/*∂v*∣ ⩽ *c*_0_, where 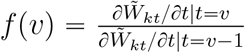 and *c*_*0*_ is a prespecified small number. We take *c*_0_ = 2*e −* 03 through the analysis.

## 3. Results

### 3.1. Fitting a generalized growth model using a time-series data

We fit the time-varying generalized growth model using the early stage outbreak series data for Wuhan, Hubei province, China, and other 41 countries with at least 2,000 confirmed cases on the date of April 9 2020. The optimal fitted model for each country or region was chosen based on the prediction error of the lowest values (Supplementary Figures 2 to 4). The corresponding empirical distributions of the parameters are shown in Table 1. The parameter *m*, the estimated mean infectious duration, ranged from 4 to 13 days, with the highest value of 13 for the epidemic in Italy. The short duration of 4 days was estimated for South Korea. The value of m reflects the duration of infectious period, which in practice could be intervened by control measures such as early diagnosis and isolation. Another parameter *η t* quantifies how rapidly the growth rate *γ*(*t*) at time *t* changed; the functional form of *η*(*t*) was estimated by pointwise estimation using data till *t* for each model respectively. The growth rates declined most rapidly in China and Thailand (median *η*: 0.86 to 0.89), and most slowly in India, Mexico, Singapore and Sweden (median *η*: 0.98 to 0.99).

**Table 1:**
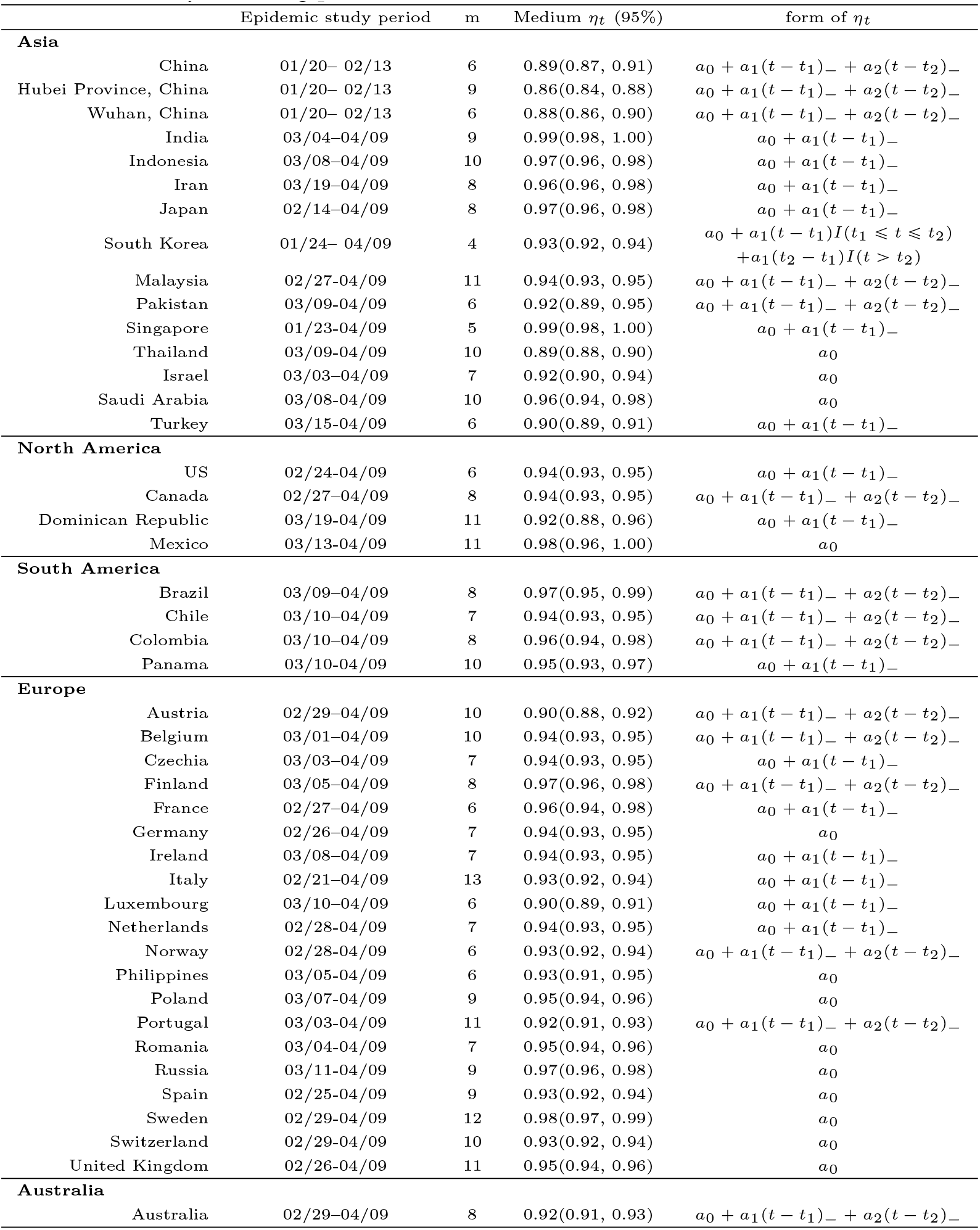
Summary of fitting parameters for the selected model in 10 countries in 2020.

| | Epidemic study period | m | Medium $\eta_t$ (95%) | form of $\eta_t$ |
| --- | --- | --- | --- | --- |
| **Asia** |  |  |  |  |
| China | 01/20– 02/13 | 6 | 0.89(0.87, 0.91) | $a_0 + a_1(t - t_1)_- + a_2(t - t_2)_-$ |
| Hubei Province, China | 01/20– 02/13 | 9 | 0.86(0.84, 0.88) | $a_0 + a_1(t - t_1)_- + a_2(t - t_2)_-$ |
| Wuhan, China | 01/20– 02/13 | 6 | 0.88(0.86, 0.90) | $a_0 + a_1(t - t_1)_- + a_2(t - t_2)_-$ |
| India | 03/04–04/09 | 9 | 0.99(0.98, 1.00) | $a_0 + a_1(t - t_1)_-$ |
| Indonesia | 03/08–04/09 | 10 | 0.97(0.96, 0.98) | $a_0 + a_1(t - t_1)_-$ |
| Iran | 03/19–04/09 | 8 | 0.96(0.96, 0.98) | $a_0 + a_1(t - t_1)_-$ |
| Japan | 02/14–04/09 | 8 | 0.97(0.96, 0.98) | $a_0 + a_1(t - t_1)_-$ |
| South Korea | 01/24– 04/09 | 4 | 0.93(0.92, 0.94) | $a_0 + a_1(t - t_1)I(t_1 \leq t \leq t_2) + a_1(t_2 - t_1)I(t > t_2)$ |
| Malaysia | 02/27–04/09 | 11 | 0.94(0.93, 0.95) | $a_0 + a_1(t - t_1)_- + a_2(t - t_2)_-$ |
| Pakistan | 03/09–04/09 | 6 | 0.92(0.89, 0.95) | $a_0 + a_1(t - t_1)_- + a_2(t - t_2)_-$ |
| Singapore | 01/23–04/09 | 5 | 0.99(0.98, 1.00) | $a_0 + a_1(t - t_1)_-$ |
| Thailand | 03/09–04/09 | 10 | 0.89(0.88, 0.90) | $a_0$ |
| Israel | 03/03–04/09 | 7 | 0.92(0.90, 0.94) | $a_0$ |
| Saudi Arabia | 03/08–04/09 | 10 | 0.96(0.94, 0.98) | $a_0$ |
| Turkey | 03/15–04/09 | 6 | 0.90(0.89, 0.91) | $a_0 + a_1(t - t_1)_-$ |
| **North America** |  |  |  |  |
| US | 02/24–04/09 | 6 | 0.94(0.93, 0.95) | $a_0 + a_1(t - t_1)_-$ |
| Canada | 02/27–04/09 | 8 | 0.94(0.93, 0.95) | $a_0 + a_1(t - t_1)_- + a_2(t - t_2)_-$ |
| Dominican Republic | 03/19–04/09 | 11 | 0.92(0.88, 0.96) | $a_0 + a_1(t - t_1)_-$ |
| Mexico | 03/13–04/09 | 11 | 0.98(0.96, 1.00) | $a_0$ |
| **South America** |  |  |  |  |
| Brazil | 03/09–04/09 | 8 | 0.97(0.95, 0.99) | $a_0 + a_1(t - t_1)_- + a_2(t - t_2)_-$ |
| Chile | 03/10–04/09 | 7 | 0.94(0.93, 0.95) | $a_0 + a_1(t - t_1)_- + a_2(t - t_2)_-$ |
| Colombia | 03/10–04/09 | 8 | 0.96(0.94, 0.98) | $a_0 + a_1(t - t_1)_- + a_2(t - t_2)_-$ |
| Panama | 03/10–04/09 | 10 | 0.95(0.93, 0.97) | $a_0 + a_1(t - t_1)_-$ |
| **Europe** |  |  |  |  |
| Austria | 02/29–04/09 | 10 | 0.90(0.88, 0.92) | $a_0 + a_1(t - t_1)_- + a_2(t - t_2)_-$ |
| Belgium | 03/01–04/09 | 10 | 0.94(0.93, 0.95) | $a_0 + a_1(t - t_1)_- + a_2(t - t_2)_-$ |
| Czechia | 03/03–04/09 | 7 | 0.94(0.93, 0.95) | $a_0 + a_1(t - t_1)_-$ |
| Finland | 03/05–04/09 | 8 | 0.97(0.96, 0.98) | $a_0 + a_1(t - t_1)_- + a_2(t - t_2)_-$ |
| France | 02/27–04/09 | 6 | 0.96(0.94, 0.98) | $a_0 + a_1(t - t_1)_-$ |
| Germany | 02/26–04/09 | 7 | 0.94(0.93, 0.95) | $a_0$ |
| Ireland | 03/08–04/09 | 7 | 0.94(0.93, 0.95) | $a_0 + a_1(t - t_1)_-$ |
| Italy | 02/21–04/09 | 13 | 0.93(0.92, 0.94) | $a_0 + a_1(t - t_1)_-$ |
| Luxembourg | 03/10–04/09 | 6 | 0.90(0.89, 0.91) | $a_0 + a_1(t - t_1)_-$ |
| Netherlands | 02/28–04/09 | 7 | 0.94(0.93, 0.95) | $a_0 + a_1(t - t_1)_-$ |
| Norway | 02/28–04/09 | 6 | 0.93(0.92, 0.94) | $a_0 + a_1(t - t_1)_- + a_2(t - t_2)_-$ |
| Philippines | 03/05–04/09 | 6 | 0.93(0.91, 0.95) | $a_0$ |
| Poland | 03/07–04/09 | 9 | 0.95(0.94, 0.96) | $a_0$ |
| Portugal | 03/03–04/09 | 11 | 0.92(0.91, 0.93) | $a_0 + a_1(t - t_1)_- + a_2(t - t_2)_-$ |
| Romania | 03/04–04/09 | 7 | 0.95(0.94, 0.96) | $a_0$ |
| Russia | 03/11–04/09 | 9 | 0.97(0.96, 0.98) | $a_0$ |
| Spain | 02/25–04/09 | 9 | 0.93(0.92, 0.94) | $a_0$ |
| Sweden | 02/29–04/09 | 12 | 0.98(0.97, 0.99) | $a_0$ |
| Switzerland | 02/29–04/09 | 10 | 0.93(0.92, 0.94) | $a_0$ |
| United Kingdom | 02/26–04/09 | 11 | 0.95(0.94, 0.96) | $a_0$ |
| **Australia** |  |  |  |  |
| Australia | 02/29–04/09 | 8 | 0.92(0.91, 0.93) | $a_0 + a_1(t - t_1)_- + a_2(t - t_2)_-$ |

### 3.2. Predictive performance of model fitting

Using the calibrated model based on m and the form of *η*(*t*), we estimated the real-time growth rates based on the number of confirmed cases at *t* 1, …, *T* by maximizing the likelihood function displayed in the Method section. The underlying growth process was lower than the constant exponential growth rate as medium *η*(*t*) was consistently estimated at less than 1(Supplementary figure 3), similar to findings of sub-exponential growth dynamics for epidemics of influenza, Ebola etc with the deceleration growth factor below 1 ^18, 19^.

For China, we used case incidence data from January 20 to February 13 as the epidemic reporting period to fit the model for trajectory predictions. The observed values fall within 95% CI of the prediction band in general, suggesting a good model fitting (Figure 1 and Supplementary Figures 8-9). We predicted that the epidemics would fade out around February 19 to 24 across the 28 provinces, about 4 weeks after the intervention starting from January 20; this is in concordance with the actual timing in which no more new cases were observed at the end of February (http://www.nhc.gov.cn). For 28 provinces in China, the estimated size was at 8553 to 9460 and 11,000 to 12,600 for Hubei province; both 95% CI estimates cover the observed figures (Table 2). For Wuhan, due to the changes in the diagnosis criteria, the predicted number of cases was not comparable after February 13. We estimated the epidemic would fade out around the end of February, which is close to the final date in which no more new cases were observed in Wuhan.

**Figure 1:**
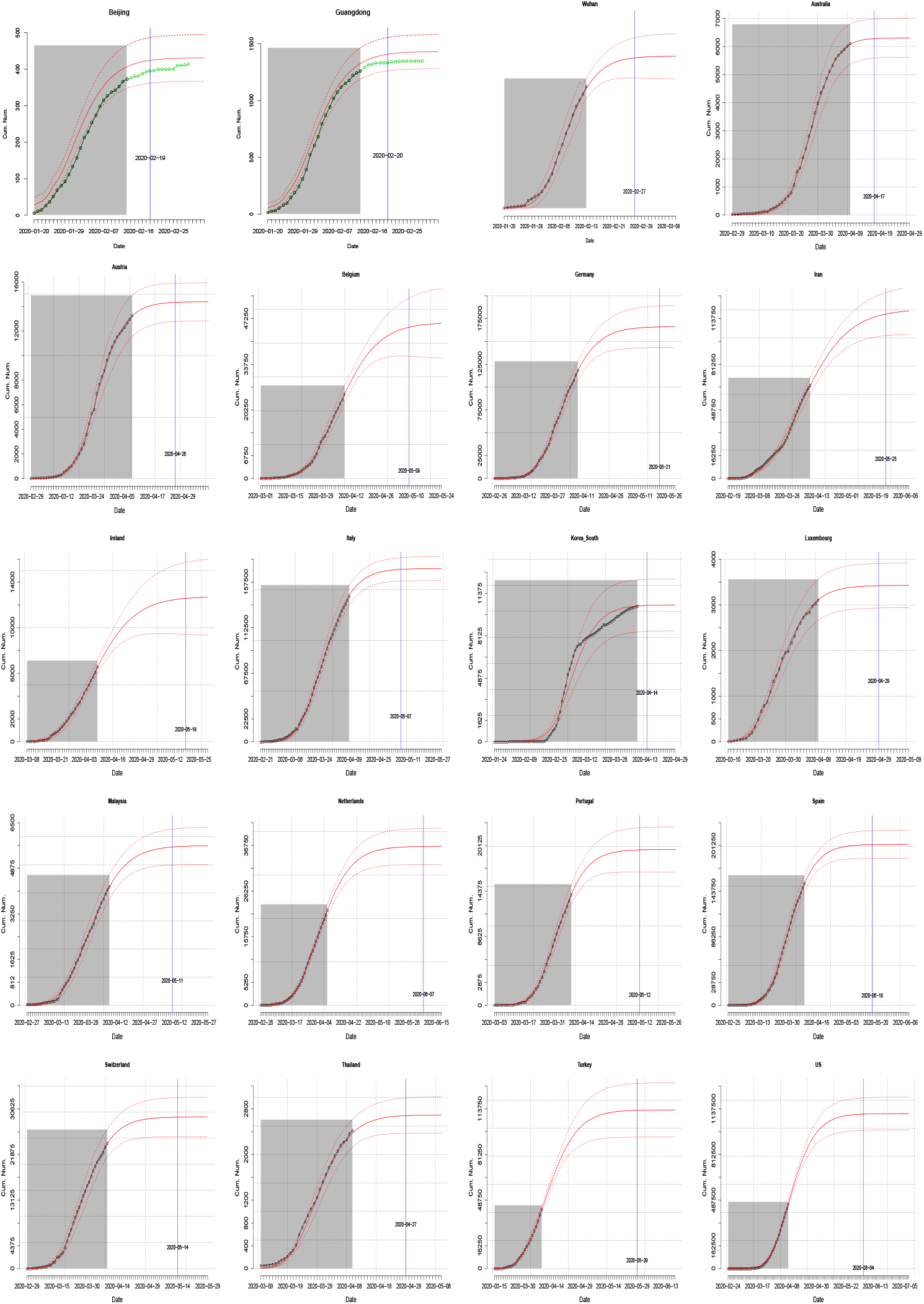
The estimated (red-solid) and observed (black-dotted) cumulative number of infectious over time *t*, as well as 95% CI (red-dashed) of the estimators. In addition, the green dots in the plot for Beijing and Guangdong in China refer to the number of reported cases which were not included for the model fitting.

**Figure 2:**
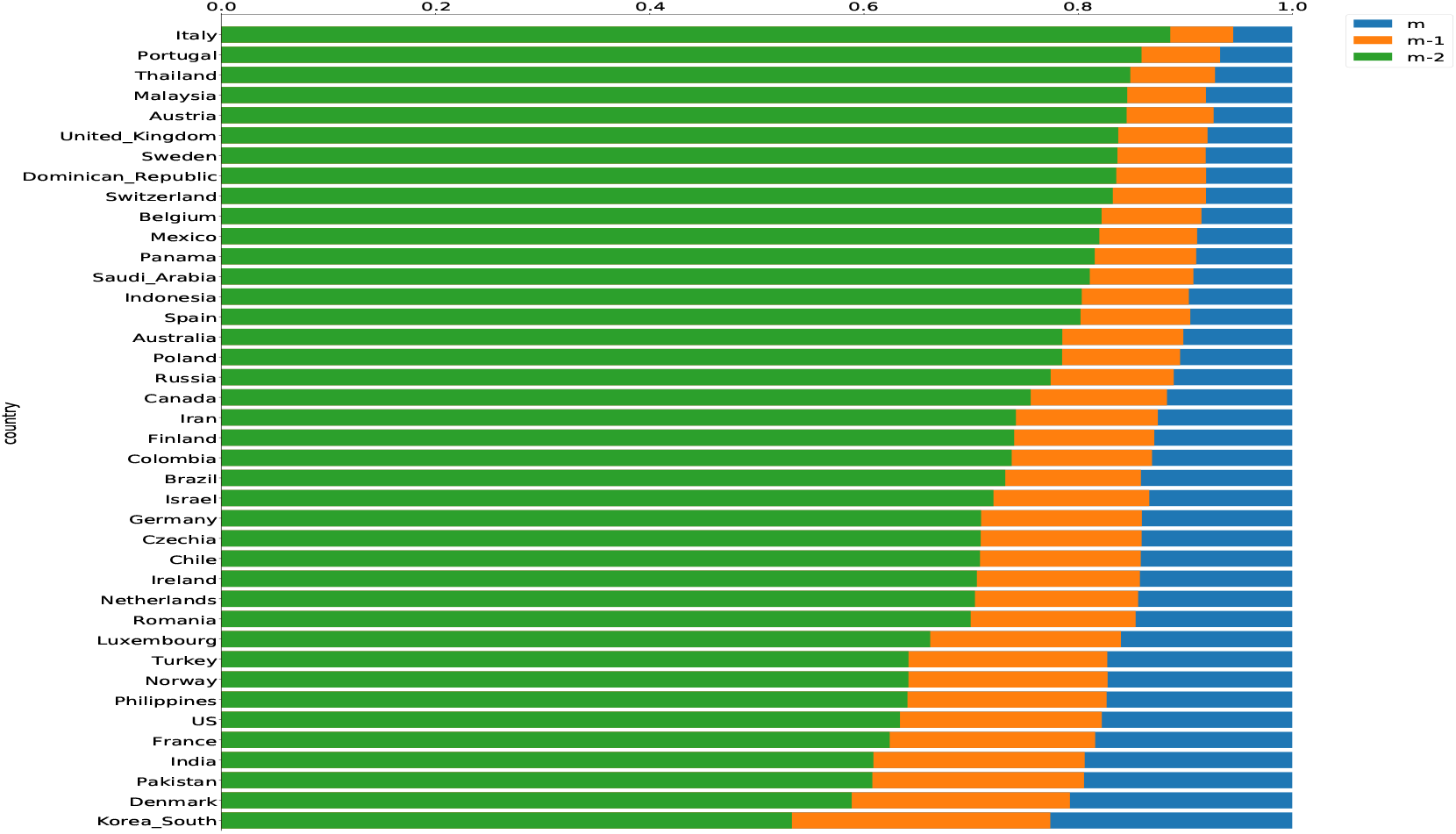
The proportion of epidemic size when the duration of infectious period *m* is reduced by 1 and 2. The reduction of infectious duration refers to shorten the time from the infection onset (symptomatic or asymptomatic) to isolation by intervention measures.

**Table 2:**
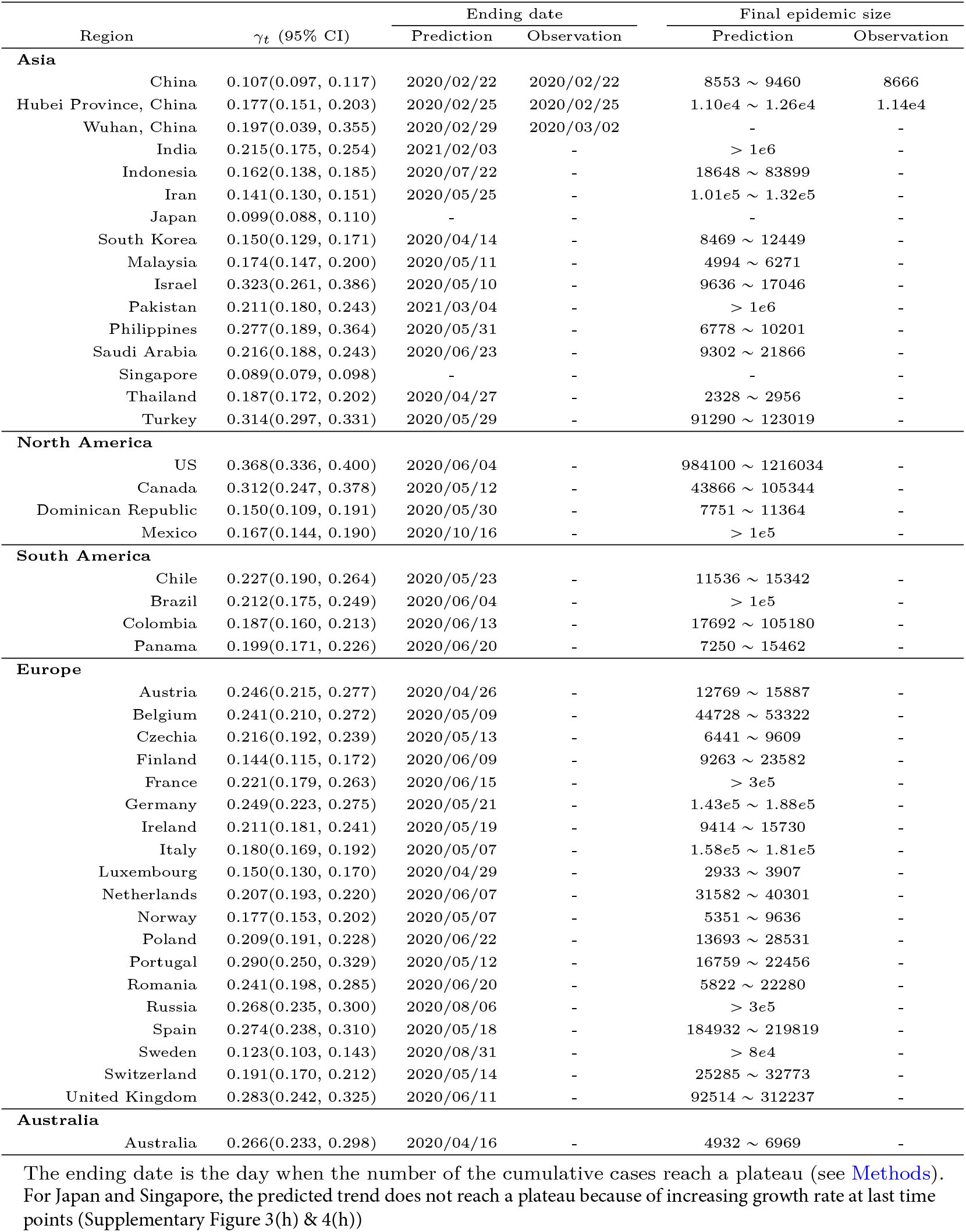
Estimation of median growth rate, final epidemic size and ending date for each country

Within each province, the observed final epidemic size was within the 95% CIs of prediction except 5 provinces, yet the prediction was still within 10% +/− flanking the upper or lower bound (Supplementary Table 1). Given the intervention measures are similar across provinces in China besides Hubei, it is not surprising that we observed the trend that the provinces with a higher estimated risk of imported cases alpha from Wuhan had an increased epidemic size. However, there were some exceptions. For example, Beijing (*α* = 28; 95%*CI* : 7 − 49) and Shanghai (*α* = 16; 4 − 28) were in the high-risk group but the final infection numbers were 286 and 261. On the contrary, Heilongjiang was in the low-risk group (*α* = 4 : 1 − 6) whereas the final infections were at 252. This implies the stronger intervention locally or compliance in densely populated municipalities (Beijing or Shanghai), than that in Heilongjiang, the north-east region far away from Wuhan. Overall our time-varying growth model provides a good fitting using time-series confirmed cases for the epidemic of COVID-19 in China.

### 3.3. Projection of the epidemic and final size in 41 countries

We projected the future growth trajectory underlying the outbreak using the daily confirmed case data from January 23, 2020 - or the date of the earliest case reported - to April 9, 2020 in 42 countries across Asia, Europe, North America, South America and Australia. Our prediction is based on the estimated parameters assuming the sustainability of intervention measures. The time-varying growth model fitting to the time-series data performed well as shown by the observed cases generally falling in the 95% CI of the prediction bound in each plot (Figure 1 and Supplementary Figures 8-12), except for South Korea in the very early phase. The predicted cases were higher than the observed number of infections from February 8 to 25, likely due to a proportion of infections being greatly understated - the hiding cases from the religious group.

We predicted the epidemic size and duration of the outbreak with the assumption of current model fitting parameters (Table 2). In Asia, the epidemic will continue until May in Philippine, Malaysia and Iran, and July in Indonesia. Strikingly, the epidemic would have the longest outbreak spread until February or March 2021 in Indian and Pakistan, with the final infections at more than a million. For Japan and Singapore, we were not able to do predictions on the epidemic sizes and duration because the estimated time-varying growth rates kept on increasing at the end of the study period – which is concordant to the big daily jump in cases observed in these two countries recently; thus incidence case data in a longer period are required for valid predictions using our model. In Europe, the epidemic will not fade out until May or June for the majority of countries. However the epidemic may last until August (Russia and Sweden). In North America, the epidemic will continue until May (Canada, Dominican Republic), June (US) and October (Mexico), and the infection are estimated to be around 1 million in the US, and 10^5^ in Mexico. In South America, the epidemic will continue until May (Chile) and June (Brazil, Colombia and Panama). Among them, Brazil has the largest estimated infections with more than 10^5^.

### 3.4. Estimation of peak time

We estimated the peak time, the day at which the incidence cases began to decline based on the estimated daily cases **d***W*_*kt*_ (see Methods). Among 41 countries, the peak time has been observed in 28 countries before or around April 9, 2020 (Table 3 & Supplementary Figures 13-15). The estimations in the above countries are concordant to the observed period during which incidence cases declined. For other countries, under the current parameters with continuity of intervention measures, we estimated the epidemic would peak in April or May 2020 for the majority of countries, except for India (July) and Pakistan (August). The predicted numbers of accumulated cases at the peak point were also consistent with the observed figures. Based on the predicted number of infections, we estimated the maximum number of ventilators for the peak requirement. For example, at least 500,000 ventilators need to be prepared for the epidemic peak in India, ∼ 100, 000 for Pakistan. Considering the death or recovery rate, the number is regarded as the upper bound in practice.

**Table 3:** The predicted or observed peak date and the corresponding cumulated size among the different countries

|  |  | Peak date |  | # Maximum cases at the peak |  | # Ventilator needed |
| --- | --- | --- | --- | --- | --- | --- |
|  |  | Prediction | Observation | Prediction | Observation | for the peak |
| Asia |  |  |  |  |  |  |
|  | India | 2020/07/18 | - | > 1e6 | - | > 5e5 |
|  | Indonesia | 2020/04/24 | - | 7804 | - | 390 ~ 780 |
|  | Iran | 2020/04/03 | 2020/03/28-04/05 | 51615 | 47226 | 2581 ~ 5162 |
|  | Israel | 2020/04/01 | 2020/03/24-04/08 | 5929 | 5717 | 296 ~ 593 |
|  | SKorea | 2020/03/04 | 2020/02/25-03/10 | 4435 | 4693 | 222 ~ 444 |
|  | Japan | - | - | - | - | - |
|  | Malaysia | 2020/03/29 | 2020/03/24-04/02 | 2469 | 2382 | 123 ~ 247 |
|  | Pakistan | 2020/08/05 | - | 1.20e6 | - | 6.01e4 ~ 1.20e5 |
|  | Philippines | 2020/04/04 | 2020/04/03-04/06 | 2914 | 3254 | 146 ~ 291 |
|  | Saudi Arabia | 2020/05/13 | - | 26803 | - | 1340 ~ 2680 |
|  | Singapore | - | - | - | - | - |
|  | Thailand | 2020/03/28 | 2020/03/22-03/29 | 1222 | 987 | 61 ~ 122 |
|  | Turkey | 2020/04/14 | 2020/04/11-04/16 | 65276 | 63144 | 3264 ~ 6528 |
| North America |  |  |  |  |  |  |
|  | US | 2020/04/09 | 2020/04/04-04/16 | 459761 | 489999 | 22988 ~ 45976 |
|  | Canada | 2020/04/08 | 2020/04/02-04/09 | 19208 | 15836 | 960 ~ 1921 |
|  | Dominican Republic | 2020/04/16 | - | 3512 | - | 176 ~ 351 |
|  | Mexico | 2020/05/23 | - | 60307 | - | 3015 ~ 6031 |
| South America |  |  |  |  |  |  |
|  | Chile | 2020/04/07 | 2020/04/03-04/16 | 5071 | 6212 | 254 ~ 507 |
|  | Brazil | 2020/05/22 | - | 3.167927e+05 | - | 15839 ~ 31679 |
|  | Colombia | 2020/04/30 | - | 8173 | - | 409 ~ 817 |
|  | Panama | 2020/04/12 | 2020/04/09-04/12 | 3083 | 2872 | 154 ~ 308 |
| Europe |  |  |  |  |  |  |
|  | Austria | 2020/03/26 | 2020/03/23-03/29 | 6641 | 6710 | 332 ~ 664 |
|  | Belgium | 2020/04/05 | 2020/04/02-04/07 | 19368 | 18875 | 968 ~ 1937 |
|  | Czechia | 2020/04/01 | 2020/03/28-04/05 | 3556 | 3586 | 178 ~ 356 |
|  | Finland | 2020/04/19 | - | 4242 | - | 212 ~ 424 |
|  | France | 2020/04/14 | 2020/04/10-04/16 | 1.60e5 | 1.21e5 | 8022 ~ 16043 |
|  | Germany | 2020/03/31 | 2020/03/24-04/08 | 72246 | 74873 | 3612 ~ 7225 |
|  | Ireland | 2020/04/06 | 2020/04/09-04/11 | 5421 | 7864 | 271 ~ 542 |
|  | Italy | 2020/03/25 | 2020/03/16-03/31 | 7.54e4 | 6.68e4 | 3771 ~ 7541 |
|  | Luxembourg | 2020/03/27 | 2020/03/21-03/31 | 1573 | 1434 | 79 ~ 157 |
|  | Netherlands | 2020/04/03 | 2020/04/02-04/10 | 15710 | 18862 | 786 ~ 1571 |
|  | Norway | 2020/03/23 | 2020/03/22-03/27 | 2659 | 3012 | 133 ~ 266 |
|  | Poland | 2020/04/12 | 2020/04/05-04/11 | 6796 | 5207 | 340 ~ 680 |
|  | Portugal | 2020/04/02 | 2020/03/31-04/10 | 8884 | 11196 | 444 ~ 888 |
|  | Romania | 2020/04/11 | 2020/04/09-04/16 | 5943 | 6424 | 297 ~ 594 |
|  | Russia | 2020/05/11 | - | 141110 | - | 7056 ~ 14111 |
|  | Spain | 2020/03/30 | 2020/03/25-04/01 | 87412 | 76795 | 4371 ~ 8741 |
|  | Sweden | 2020/05/09 | - | 37389 | - | 1869 ~ 3739 |
|  | Switzerland | 2020/03/27 | 2020/03/23-03/30 | 12516 | 12392 | 626 ~ 1252 |
|  | United Kingdom | 2020/04/13 | 2020/04/05-04/14 | 86715 | 70811 | 4336 ~ 8672 |
| Australia |  |  |  |  |  |  |
|  | Australia | 2020/03/28 | 2020/03/25-03/30 | 3482 | 3384 | 174 ~ 348 |

\* Peak date has been observed. Due to the fluctuation of the daily reported cases, the observed peak interval was listed instead of the exact peak date.
The number of maximum cases at the peak is the accumulated number of cases at the peak period.
The number of ventilator for the peak requirement is calculated based on the predicted infection. We assume 5 – 10% of the infected patients need the ventilator<sup>22</sup>.

### 3.5. Impact of the reduction of the infectious period

Shortening the time from the infection onset (symptomatic or asymptomatic) to isolation, termed as ‘infectious period’ in this paper, is vital as it will reduce transmission. The control strategies could rely on social distancing, earlier isolation of cases and population-based testing to identify the presymptomatic or asymptomatic cases. Here we evaluated the impact of shortening the infectious period on the epidemics, showings the epidemic curve for each country, assumed to be 1 or 2 days shorter in the duration of infectivity infectious period (Supplementary Figures 13-15). A reduction of the infectious period by 1 or 2 days would have a negligible effect on the peak time, as the estimation was essentially the same. The reduction of the infectious period by 1 or 2 days would lead to a significantly flatted epidemic curve across all the countries. For example, the 2-days decrease in the infectious period would result in a significant reduction of the final epidemic size in India (31.1%), Pakistan (39.3%), Russia (22.6%), Brazil (26.8%), and Mexico(18.0%).

### 3.6. Estimation of real-time effective reproduction number R_t_

We quantified the real-time reproduction number *R*_*t*_ (see Method section)^17, 20, 21^, based on the estimation parameters of growth rates *γ* and the infectious period *m*. The *R*_*t*_ exhibited a declining trend with time and variability in the estimates across countries (Table 4). The uncertainty of Rt was largest in the first week and gradually became smaller with time (Table 4). In the first week, 28 countries displayed *R*_*t*_ below 5 while 14 exhibited large *R*_*t*_ above 5. Italy and Spain displayed the highest *R*_*t*_ at 12.36, and 9.26 respectively. *R*_*t*_ estimations become closer ranging from 0.95 (Singapore) to 5.54 (Spain) in the second week. During the first month of the epidemic period, all countries displayed a declined trend towards 1 in the epidemic period with varying deceleration rates, revealing the impacts of the intervention strategies. In the fourth week, 14 countries exhibited *R*_*t*_ below 1, the highest in UK (2.62) and the US (2.19). For Singapore and Japan, although both displayed the declining trend in the first month, there is an inclining trend starting from April, implying the intervention could not be effective in the late stage (Supplementary Figure 16). *R*_*t*_ displayed the fastest deceleration in China with the most pronounced changes during the third to fourth weeks; this may reveal the significant impact of intervention measures implemented since January 20, 2020 in the first week.

**Table 4:** Estimated Medium *R*_*t*_ (95%CI) in the subsequent week

|  |  | First week | Second week | Third week | Fourth week |
| --- | --- | --- | --- | --- | --- |
| **Asia** | China | 1.64(0.98, 2.30) | 1.34(1.21, 1.48) | 0.59(0.55, 0.63) | 0.26(0.24, 0.28) |
|  | Hubei Province, China | 3.11(2.58, 3.64) | 2.05(1.86, 2.23) | 0.73(0.68, 0.77) | 0.26(0.23, 0.29) |
|  | Wuhan, China | 2.50(1.56, 3.44) | 1.51(0.83, 2.19) | 0.64(0.40, 0.88) | 0.27(0.17, 0.37) |
|  | India | 2.34(1.31, 3.38) | 1.23(0.83, 1.63) | 1.12(0.81, 1.42) | 1.02(0.71, 1.32) |
|  | Indonesia | 4.49(3.66, 5.31) | 1.77(1.30, 2.23) | 1.24(0.98, 1.51) | 0.99(0.87, 1.11) |
|  | Iran | 3.56(2.39, 4.73) | 2.01(1.78, 2.23) | 1.37(1.21, 1.53) | 1.05(0.94, 1.16) |
|  | Israel | 7.70(3.86, 11.55) | 4.26(2.79, 5.73) | 2.36(1.88, 2.83) | 1.30(1.16, 1.45) |
|  | Japan | 4.36(3.21, 5.50) | 1.89(1.55, 2.22) | 1.02(0.90, 1.14) | 0.68(0.60, 0.76) |
|  | SKorea | 1.52(0.99, 2.06) | 1.42(0.99, 1.84) | 1.32(0.98, 1.65) | 1.22(0.96, 1.49) |
|  | Malaysia | 5.77(4.81, 6.72) | 3.39(2.77, 4.02) | 2.19(1.66, 2.72) | 1.46(1.18, 1.74) |
|  | Pakistan | 4.90(3.58, 6.22) | 2.32(1.82, 2.83) | 1.02(0.86, 1.18) | 0.71(0.60, 0.82) |
|  | Philippines | 4.18(1.20, 7.17) | 2.59(1.31, 3.87) | 1.60(1.12, 2.09) | 0.99(0.83, 1.16) |
|  | Saudi Arabia | 2.91(1.55, 4.27) | 2.14(1.53, 2.75) | 1.57(1.33, 1.80) | 1.15(0.97, 1.33) |
|  | Singapore | 1.38(1.05, 1.72) | 0.95(0.77, 1.14) | 0.70(0.59, 0.81) | 0.54(0.47, 0.61) |
|  | Thailand | 6.03(4.55, 7.50) | 2.99(2.60, 3.37) | 1.48(1.38, 1.58) | 0.73(0.63, 0.84) |
|  | Turkey | 5.53(4.10, 6.95) | 1.97(1.85, 2.10) | 1.08(0.95, 1.21) | - |
| **North America** | US | 3.03(2.53, 3.54) | 3.69(3.13, 4.26) | 3.24(2.68, 3.79) | 2.19(1.91, 2.47) |
|  | Canada | 2.92(2.50, 3.34) | 2.25(1.60, 2.91) | 2.45(1.78, 3.13) | 2.00(1.58, 2.41) |
|  | Dominican Republic | 3.91(2.87, 4.94) | 1.16(0.52, 1.80) | 0.84(0.60, 1.08) | - |
|  | Mexico | 2.78(1.80, 3.76) | 2.45(2.00, 2.90) | 2.16(1.99, 2.32) | 1.90(1.68, 2.11) |
| **South America** | Chile | 5.29(4.03, 6.54) | 2.33(1.78, 2.89) | 1.31(1.09, 1.53) | 0.85(0.76, 0.95) |
|  | Brazil | 6.91(4.55, 9.27) | 4.36(3.30, 5.41) | 1.84(1.56, 2.12) | 1.54(1.38, 1.71) |
|  | Colombia | 5.37(3.82, 6.92) | 2.55(1.90, 3.19) | 1.34(1.13, 1.55) | 1.07(0.90, 1.24) |
|  | Panama | 4.89(2.67, 7.11) | 2.03(1.51, 2.55) | 1.41(1.07, 1.76) | 1.00(0.70, 1.30) |
| **Europe** | Austria | 4.91(3.63, 6.19) | 3.91(2.80, 5.01) | 2.75(2.14, 3.36) | 1.38(1.17, 1.58) |
|  | Belgium | 6.61(4.95, 8.28) | 4.27(3.36, 5.19) | 2.67(2.26, 3.08) | 1.76(1.59, 1.92) |
|  | Czechia | 4.07(2.81, 5.33) | 2.33(2.07, 2.59) | 1.47(1.28, 1.67) | 0.93(0.76, 1.09) |
|  | Finland | 7.76(5.23, 10.28) | 2.50(1.23, 3.78) | 1.38(0.80, 1.97) | 1.10(0.72, 1.48) |
|  | France | 4.29(3.39, 5.19) | 1.80(1.06, 2.54) | 1.26(0.83, 1.69) | 0.95(0.71, 1.18) |
|  | Germany | 5.52(4.16, 6.88) | 3.48(2.83, 4.13) | 2.19(1.91, 2.48) | 1.38(1.27, 1.49) |
|  | Ireland | 4.08(2.88, 5.27) | 1.77(1.36, 2.18) | 1.15(0.92, 1.38) | 0.76(0.63, 0.90) |
|  | Italy | 12.36(10.99, 13.74) | 5.14(4.52, 5.76) | 3.11(2.77, 3.46) | 1.91(1.71, 2.10) |
|  | Luxembourg | 4.05(2.48, 5.63) | 1.71(1.47, 1.94) | 0.82(0.70, 0.94) | 0.40(0.33, 0.47) |
|  | Netherlands | 4.39(3.01, 5.77) | 2.54(2.38, 2.70) | 1.68(1.57, 1.79) | 1.13(1.04, 1.22) |
|  | Norway | 3.89(3.09, 4.70) | 2.43(2.09, 2.77) | 1.38(1.09, 1.66) | 0.75(0.62, 0.89) |
|  | Poland | 3.45(2.85, 4.05) | 2.46(2.15, 2.77) | 1.75(1.61, 1.90) | 1.25(1.17, 1.33) |
|  | Portugal | 5.87(4.44, 7.30) | 4.30(3.38, 5.22) | 3.24(2.74, 3.74) | 1.83(1.68, 1.99) |
|  | Romania | 3.42(2.06, 4.78) | 2.40(1.73, 3.08) | 1.69(1.38, 1.99) | 1.19(1.06, 1.31) |
|  | Russia | 3.41(2.61, 4.22) | 2.71(2.28, 3.14) | 2.15(1.97, 2.33) | 1.70(1.62, 1.79) |
|  | Spain | 9.26(6.73, 11.79) | 5.54(4.34, 6.73) | 3.31(2.78, 3.84) | 1.98(1.76, 2.20) |
|  | Sweden | 2.39(1.64, 3.14) | 2.07(1.56, 2.58) | 1.79(1.47, 2.12) | 1.55(1.35, 1.75) |
|  | Switzerland | 6.39(4.64, 8.15) | 3.77(3.05, 4.49) | 2.22(1.95, 2.50) | 1.31(1.18, 1.44) |
|  | United Kingdom | 7.46(5.18, 9.74) | 5.26(4.00, 6.52) | 3.71(3.06, 4.36) | 2.62(2.30, 2.93) |
| **Australia** | Australia | 3.72(3.41, 4.03) | 2.48(2.16, 2.79) | 2.16(1.90, 2.41) | 1.43(1.30, 1.57) |

## 4. Discussions

Here we predicted that the COVID-19 pandemic will persistently spread over many countries and will last on average 5 to 6 months since January 2020 for the current wave. We estimated, given no further intervention measures, that India would emerge to be a new epidemic center, as well as Pakistan, Brazil, Russia and Mexico. Effective intervention measures by reducing the infectious period would result in an 50% reduction in the final epidemic sizes. *R*_*t*_ had a declining trend in almost all countries, revealing the impact of the intervention measures i.e. social distancing.

Our real-time estimation framework can yield a reliable prediction of the final epidemic size using the data from the early phase of the epidemic. We evaluated the performance of our model using incident case data from January 20 to February 13, 2020 in China. We predicted the epidemic would fade out on February 19 to 24 across 28 provinces in China with a one-week lag in Wuhan; the epidemic size was estimated at 8,500 to 9,500, and 11,000 to 12,600 in 28 provinces and Hubei respectively. The outbreak size and epidemic duration estimated are found to be highly consistent with the observed figures (https://www.who.int). The prediction of the final epidemic size based on the model that assumes early exponential growth could tend to overestimate the epidemic size, which has been shown in the previous studies^10,23-28^

We estimated that the most affected countries in the next wave of the pandemic will be India, Pakistan, Brazil, Mexico as well as Russia. All these countries are currently undergoing regional or national lockdown except Mexico. Given that thousands of ventilators are needed for the peak around May to July, it is immediately important for these countries to prepare the ICU beds. In Europe, most countries have peaked in the curve, and the final outbreak size was estimated to exceed 10^5^ in Italy, France, UK, Germany and Spain. For the United States, the estimated final epidemic size will surpass 1 million, peaking around July 2020. A recent report calculated that 81% of British and the United States populations would be infected, with approximately a half and 2.2 million deaths respectively^29^. The high value is based on the assumption of *R*_0_ at 2.4, while our results demonstrate that real-time *R*_*t*_ decelerated under interventions. Thus, their estimates are likely to be the higher bound of the true value. In Asia, besides China, South Korea and Iran, Thailand and Malaysia have peaked in March. Indonesia and Saudi Arabia would peak in April. Singapore and Japan recently implemented lockdown and more data will be needed for the epidemic prediction. Given the case-fatality rate at 2.3%^6^, we estimated around 23,000 deaths in India and Pakistan, and around 2,300 in Mexico and Brazil, and 6,900 in Russia.

This is the first study to compare country-specific temporal *R*_*t*_. It is natural to expect a declining trend of *R*_*t*_ to 1 owing to stochastic effects for epidemics governed by subexponential growth^18, 30^, while a faster decline may suggest a larger impact of intervention measures or behavior changes31. A majority of countries displayed *R*_*t*_ below 5 in the first week of the epidemic period. Several countries exhibited large *R*_*t*_, ie. Italy or Spain, suggesting a rapid increase of cases at the beginning or a large variation in the under-reporting rates in the early epidemic phase. For Wuhan, *R*_*t*_ declined from the median 2.5 to 0.3 within a month, which is compatible with the *R*_0_ estimation in a range of 2.2 to 3.6 in Wuhan^2, 10, 12, 13, 16, 23, 31^, and below 1 after interventions^32^. All countries displayed a declined trend towards 1 in the epidemic period with varying deceleration rates, revealing the variation in the impact of the intervention strategies. For further research on the intervention effectiveness, individual data, as well as a series of intervention measures for each country, may be required to quantify the effects in detail.

Our study showed that by shortening the infectious period by two days, we could effectively reduce the final epidemic size by up to 50%. What strategies can effectively reduce the infectious period? Usually, it can be done - similar to the other coronaviruses - by early detection and isolation of symptomatic patients and tracing of close contacts. However, the presymptomatic transmission has been reported in many countries such as China, the US, and Singapore^33–36^, etc. The existence of presymptomatic or asymptomatic transmission would present difficult challenges for disease control, which underscores the importance of social distancing. For instance, at the onset of the epidemic, the spread has been well controlled in Singapore, accounted for by a range of intervention measures, such as contact tracing and quarantining, that were instituted from January 23-the onset of the first case in Singapore^37^. However, the recent increase in cases in Singapore, initially starting from the imported cases followed by the outbreak in dormitories of immigration workers, led to its circuit period from April 7 to May 4. Quarantine measures for the infected regions or at the national scale, either a complete lockdown as in China or partial lockdown such as in Europe, are effective in flattening the curve^38^. Besides the quarantine measures, South Korea was also successful in suppressing the outbreak, attributed to the rapid measures to perform large-scale diagnostic testing for the public for case isolation^39^.

During the early phase of the epidemic, we forecasted the epidemic size as a function of the time-varying growth model, similar but more flexible to the previous approaches to model sub-exponential growth dynamics^18–21, 40^, without relying on any epidemiological parameter assumptions. The traditional transmission epidemiological model is defined through a susceptible, exposure, infectious, removed (SEIR) scheme, which requires epidemiological parameters from detailed case studies^2, 10, 31, 41^. Furthermore, given the variation of the transmissibility, mostly due to the intervention strategies implemented and behavioral changes in the population, it is desirable to quantify the dynamics of *R*_*t*_ over time^20,42-44^. Several studies attempted to forecast the number of epidemics for COVID-19 in China using the constant growth rate^27, 45, 46^, which is not the case in our model of nonmonotonic behavior of the growth rate. Our model, compared to previous studies, has greater flexibility in a data-mining manner to fit and predict future trajectories.

In summary, we compared the epidemic trajectory, characterized dynamic *R*_*t*_ in 42 countries, and predicted that the new epidemic centers will continue to emerge in the next wave. Meanwhile, we highlighted the importance of various effective interventions in flattening the epidemic curve, such as social distancing, to shorten the infectious period. By carefully characterizing the shape of the epidemic growth phase, we believe our study represents a significant step in modeling real-time transmission and providing accurate forecasts of epidemics.

Our study has several limitations. Firstly, the projection of the temporal trend of an outbreak using the early-stage dataset could be dramatically influenced by the changes in the intervention strategies later on. For example, using data up to March 15, our previous analysis overestimated the epidemic size in U.S as the social distance measures (i.e. quarantine) were implemented starting from March 21. The performance of models used in this work will be continuously improved with data coming in from an ongoing outbreak, thus real-time estimates of key epidemiological parameters can be available before the epidemic fully ends. Secondly, the number of infections estimated might not be comparable across the countries; for example, the number of infections in Germany is not comparable to the number of infections in Italy or China, as the latter did not perform large-scale population-based testing and thus the cases could be more severe. Lastly, the analyses are highly reliant on the reporting criteria and quality of the data. The under-reporting of infection is likely a common scenario in the majority of countries. For example, a recent study showed that the reported number of confirmed positive cases was 50-85-fold lower than the actual number of infections in 3330 people in Santa Clara County, US^47^. A more realistic and comprehensive analysis could be performed that includes accurate epidemic data and information. In current imperfect situation, our model could still be used for more advanced analyses, including estimations of the epidemic size, peak points and dynamic *R*_*t*_.

## Data Availability

The data used in this article is publicly available, see the data links in the article for details.

## Acknowledgments

The research were partially supported by National Natural Science Foundation of China (Nos. 11931014 and 11829101) and Fundamental Research Funds for the Central Universities (No. JBK1806002) of China.

